# Prevalence and Determinants of E-Cigarette Use Among Diploma Students in a Vocational College: A Cross-Sectional Study

**DOI:** 10.1101/2024.09.23.24314252

**Authors:** Siti Munisah Mohd Shoaib, Norliza Ahmad, Aidalina Mahmud

## Abstract

**Introduction:** The prevalence of e-cigarette use is increasing globally, particularly among young adults which can predispose them to various health risks. This study aimed to determine the prevalence and factors associated with e-cigarette use among diploma students in a vocational college in a state in Malaysia.

**Methodology:** A cross-sectional study using probability proportionate to size sampling was conducted among 700 diploma students at a vocational college in Malaysia. A validated, self-administered questionnaire was distributed through the online method from April to May 2024. Bivariate analysis was done using Pearson’s chi-square test or Fisher’s exact and simple logistic regression. Multivariable analysis was performed using multiple logistic regression for variables with a p<0.25 in the bivariate analysis. A value of p<0.05 was considered statistically significant, with a 95% confidence interval.

**Results:** The response rate was 87.7% with the prevalence of e-cigarette use was 29.0%. Factors significantly associated with e-cigarette use included male (aOR = 5.2, 95% CI: 2.7- 10.1), other races (aOR = 83.1, 95% CI: 2.2-3146.3), perceived e-cigarette aids in quit smoking (aOR = 1.6, 95% CI: 1.2-2.1), perceived e-cigarette does not contain the toxic chemicals found in conventional cigarette (aOR = 1.4, 95% CI: 1.0-2.0), having close friends who use conventional cigarette (aOR = 2.1, 95% CI: 1.0-4.1) or e-cigarette (aOR = 8.0, 95% CI: 2.3- 28.1), e-cigarette exposure on television (aOR = 2.1, 95% CI: 1.0-4.2), positive attitude towards e-cigarette (aOR = 1.2, 95% CI: 1.1-1.2), higher willingness to use (aOR = 1.2, 95% CI: 1.0-1.3), and higher intention to use (aOR = 1.4, 95% CI: 1.2-1.5).

**Conclusion:** Factors associated with e-cigarette use among diploma students included being male, other races (Bumiputera Sabah and Sarawak), positive health risks perceptions, peer influence, and exposure to e-cigarettes on television. Targeted interventions addressing these factors may be more effective in changing social norms and reducing e-cigarette use among this population.

## Introduction

E-cigarettes are devices that heat a liquid, typically containing nicotine, flavorings, and other substances to produce an aerosol that is inhaled by the user. E-cigarette use poses significant health risks, particularly to young adults, youth, and pregnant women (1). Currently, there are rising concerns about the increasing trend of e-cigarette use. Previous studies conducted among college and university students aged 18 to 26 years have reported a wide range of prevalence rates. A study in Malaysia in 2016 reported a prevalence of 20.4%, whereas other studies done in the U.S. in 2019, Thailand, and Malaysia in 2020 reported a prevalence of 24.8%, 18.1%, and 12.4% respectively (2–5). A study conducted among vocational students in Thailand in 2019 reported an even higher prevalence of current e-cigarette use (28.7%), with the highest percentage seen among respondents aged 20 years and older (30.8%) (6).

Nicotine contained in e-cigarette liquid may impair brain cognition and development, increase impulsive behavior, and negatively impact psychosocial health in young adults (7–9). Other chemicals in e-cigarettes, such as flavorings, diacetyl, propylene glycol (PG), carbon monoxide, formaldehyde, and polycyclic aromatic compounds, are also very harmful to health (7,10). In fact, respiratory problems, such as asthma, chronic obstructive pulmonary disease, eosinophilic pneumonia, epiglottitis, bronchitis, and acute respiratory distress, have been linked to e-cigarette use (7). Additionally, e-cigarette or vaping use-associated lung injury (EVALI) has been reported among e-cigarette users, which is caused by tetrahydrocannabinol (THC) and vitamin E acetate present in some e-cigarettes (11,12).

Other harmful health risks of e-cigarette use include accidental nicotine overdose, increased risk of cardiopulmonary diseases, subsequent illicit drug use, serving as a gateway to conventional cigarettes that may result in dual use of both types of cigarettes, and physical injuries from battery failures and explosions (7,13). Small liquid droplets produced by e-cigarette aerosols have also been detected in the environment and on surfaces, potentially causing second-hand and third-hand exposure to people nearby (14,15). Among those exposed to second-hand and third-hand e-cigarette aerosols smoker, respiratory problems (especially among those with existing airway conditions such as asthma), DNA damage, immediate modifications in respiratory mechanics and exhaled inflammatory biomarkers, and an increased risk of cardiovascular disease have been observed (14,16–18).

One of the intrapersonal factors of e-cigarette use is having lower levels of knowledge about the potential negative health outcomes of e-cigarette use (4). Other intrapersonal factors, such as poor health risk perceptions and poor mental health perceptions, have also been identified among e-cigarette users (5,19,20). E-cigarette use is also influenced by family members and peers, exposure to printed or online advertisements, the availability and affordability of e-cigarettes, positive attitudes towards e-cigarettes, higher intention and higher willingness to use, as well as engagement in high-risk behaviors have also been found to be associated with the use of e-cigarettes (5,6,19,21–28). In Malaysia, the National Health Morbidity Survey (NHMS) 2019 indicated that the highest prevalence was among the age group of 20 to 24 years (14.7%) (29). However, studies among students at vocational colleges on e-cigarette use are scarce. Hence, this study aimed to identify factors associated with e-cigarette use among young adults currently studying at a vocational college in Selangor, Malaysia.

## Materials and Methods

### Study design and population

A cross-sectional study was conducted among diploma students at a vocational college in Selangor, Malaysia, from October 2023 to July 2024. Selangor, a state adjacent to Kuala Lumpur, the capital of Malaysia, was selected as the study site due to its high prevalence of e-cigarette use, as reported in the NHMS 2019. In addition, it houses the highest number of higher education institutions, with a total of 397,366 enrolments in 2022 (29,30). Inclusion criteria included students currently enrolled in semester two or beyond, and young adults aged 18 to 26 years. Students who were on leave, not attending classes during the data collection period, or were suspended by the university were excluded.

### Sample size and sampling

The required sample size was 700 participants, at 80% power, 95% confidence level, and 20% non-response rate. The sample size was determined using the two-population proportion formula, based on e-cigarette use and monthly family income from a previous study (5). Stratified random sampling with probability proportionate to size method was employed in this study. All 12 diploma programs (regarded as strata) in the selected college were included. Probability proportionate to size sampling was applied to each diploma course based on the calculated sample size. Finally, systematic random sampling was used to select eligible participants from each diploma course.

### Study instrument

This study used a set of validated questionnaires adapted from previous research (20,24–26,28,31,32). It consisted of 12 sections. Part A of the questionnaire was regarding the sociodemographic characteristics and academic performance of the respondents. Part B of the questionnaire was regarding conventional cigarette and e-cigarette use profiles which were assessed by whether or not the respondent had smoked a conventional cigarette or use e-cigarette in the past 30 days with three response options (31). E-cigarette users were then categorised into “never user” and “e-cigarette user” accordingly. Part C was regarding reasons for using e-cigarettes which were answered by those respondents who are occasional e-cigarette users and past month e-cigarette users only with six options were given (2). The answer options for the questions will be “Yes” and “No”. Part D was regarding knowledge of e-cigarettes with eight statements (26). The answer options were “Yes”, “No”, and “Don’t know”. Those who answered “Don’t know” were interpreted as wrong answers which were scored zero, while the correct answer was scored one. The minimum score was 0 and the maximum score was 8. A higher score will reflect good knowledge of e-cigarettes.

Part E consisted of two constructs on perceptions. The first construct consisted of five statements to assess health risk perceptions on e-cigarette (25). It was scored using a 5-point Likert scale (1-totally disagree, 2-disagree, 3-unsure, 4-agree, 5-totally agree). The minimum score was 5 and the maximum score was 25. A lower score reflected that the respondents have a strong belief in the health risks of e-cigarettes. The second construct was on mental health perceptions (20). It consisted of four questions on perceived anxiety, depression, loneliness, and stress symptoms during the past month. The questions were measured using a 5-point Likert scale (1-never, 2-rarely, 3-sometimes, 4-often, 5-always). The minimum score was 4 and the maximum score was 20. A higher score reflected poor mental health perceptions.

Part F was regarding social influences with two statements about behaviour of conventional cigarette and e-cigarette use among family members and another two statements about behaviour of conventional cigarette and e-cigarette use among close friend (25). The answer option was “Yes” and “No”. Part G was regarding advertising media influences based on the exposure to e-cigarette advertisements (24). A statement with six options was given with “Yes” and “No” answer options displayed for each option.

Part H was regarding e-cigarette availability and affordability (25). The availability was explored by whether the respondents think it is difficult to get e-cigarettes. Meanwhile, the affordability was explored by whether the respondents think the price of e-cigarettes is affordable. The answer options for both questions were “Yes” and “No”. Part I, part J and K was regarding attitude towards e-cigarettes, willingness to use, and intention to use respectively (32). Attitude towards e-cigarette consisted of seven statements while willingness to use consisted of three statements and was scored using a 5-point Likert scale (1-totally disagree, 2- disagree, 3-unsure, 4-agree, 5-totally agree). Meanwhile, intention to use consisted of three questions and scored using a 5-point Likert scale (1-very unlikely, 2-unlikely, 3-unsure, 4- likely, 5-very likely). A higher score reflected a more positive attitude towards e-cigarettes, a higher willingness to use e-cigarettes, and a higher intention to use e-cigarettes. Part L was regarding high-risk behaviours with three statements on alcohol use, illegal drug use, and unprotected sex with single or multiple sexual partners (28). The answer options were “Yes” and “No”.

Pre-testing was conducted among 30 students from a diploma course to assess the stability of the questionnaire. Data from this pre-test was excluded from the final analysis. The internal consistency of the questionnaire was evaluated using Cronbach’s alpha, with values ranging from 0.70 to 0.96, indicating good to excellent reliability (33). The Cronbach’s alpha values for each construct are summarized in Table 1.

**Table 1:** Summary of the Cronbach’s alpha index value.

| Domain | Number of Items | Cronbach's Alpha |
| --- | --- | --- |
| Health risk perception | 5 | 0.70 |
| Mental health perceptions | 6 | 0.77 |
| Attitude towards e-cigarette | 7 | 0.89 |
| Willingness to use e-cigarette | 3 | 0.74 |
| Intention to use e-cigarette | 3 | 0.96 |

### Data collection

Data was collected online using Google Forms, which were distributed to respondents through the WhatsApp application. Data collection took place from April to May 2024. Prior to data collection, a brief presentation detailing the study’s objectives and methodology was given to the head of the department. Respondent eligibility was assessed based on the inclusion and exclusion criteria.

### Statistical analysis

Data were analyzed using IBM SPSS version 29.0. Prior to analysis, the data sets were examined for out-of-range values and assessed for normality. Continuous independent variables were presented as mean ± standard deviation for normally distributed data and as median (interquartile range) for non-normally distributed data. Continuous independent variables, such as respondents’ age, monthly family income, and GPA, were categorized accordingly. Categorical variables were presented as frequencies and percentages. Bivariate analysis was conducted using the chi-square test for categorical independent variables and simple logistic regression for continuous independent variables to assess associations with e-cigarette use.

Results of bivariate analyses with p-value <0.25 were further tested for multicollinearity and interactions before being analyzed using multiple logistic regression to determine the factors associated with e-cigarette use (34). Three regression methods were employed: the “Enter” method, the “Forward LR” method, and the “Backward LR” method. The most parsimonious model that best fits the data was selected. Results were reported as crude and adjusted odds ratios, with statistical significance set at p < 0.05.

### Ethics approval and consent to participate

Ethical clearance was obtained from the Ethics Committee for Research Involving Human Subjects (JKEUPM), Universiti Putra Malaysia (JKEUPM-2024-193), in accordance with ethical standards. Approval from the Polytechnic and Community College Education Department, Ministry of Higher Education, was obtained prior to the pre-test and data collection. Online consent was obtained from all respondents via Google Forms. All information and details collected from the respondents were kept anonymous and transferred to a password-protected database linked to student identification created for this study. Access to this database was restricted to the investigators and will be maintained for seven years after the completion of the study, after which it will be deleted.

## Results

Out of 700 randomly selected respondents invited from the sampling list, 614 students participated, resulting in an overall response rate of 87.7%. The prevalence of e-cigarette use in this study was 29.0%. Most respondents were aged between 18 and 20 years (73.0%), male (55.4%), of Malay ethnicity (87.6%), had a household average monthly income in the lower 40% (B40) income group (83.9%), and achieved a Grade Point Average (GPA) grade of B (76.2%). Table 2 presents the distribution of respondents according to sociodemographic characteristics and academic performance.

**Table 2:** Distribution of respondents according to sociodemographic characteristics and academic performance (n=614)

| Respondent characteristics | Mean±SD / Median (IQR) | Frequency (n) | Percentage (%) |
| --- | --- | --- | --- |
| **Age** | 20 (2) |  |  |
| 18 – 20 years old |  | 448 | 73.0 |
| 21 – 25 years old |  | 166 | 27.0 |
| **Sex** |  |  |  |
| Male |  | 340 | 55.4 |
| Female |  | 274 | 44.6 |
| **Race** |  |  |  |
| Malay |  | 538 | 87.6 |
| Chinese |  | 14 | 2.3 |
| Indian |  | 53 | 8.6 |
| Others |  | 9 | 1.5 |
| **Monthly family income** | 3000 (3000) |  |  |
| B40 |  | 515 | 83.9 |
| M40 |  | 78 | 12.7 |
| T20 |  | 21 | 3.4 |
| **GPA result** | 3.21±0.41 |  |  |
| A |  | 93 | 15.1 |
| B |  | 468 | 76.2 |
| C |  | 53 | 8.7 |

The overall mean score for knowledge was 4.60±1.64 out of 8. Respondents were almost equally distributed between those scoring 50% or below (45.3%) and those scoring above 50% (54.7%). Among the respondents, only 10 (1.6%) answered all the questions correctly, while 13 (2.1%) scored zero. Most respondents (88.6%) correctly identified that e-cigarettes represent a new form of addiction in society. Meanwhile, 75.2% incorrectly believed that e-cigarettes do not contain tar. Details on the distribution of knowledge among respondents are summarized in Table 3.

**Table 3:** Distribution of respondents according to knowledge of e-cigarettes (n=614)

| Knowledge of e-cigarette | Correct answer | Incorrect answer |
| --- | --- | --- |
|  | Frequency (%) | Frequency (%) |
| E-cigarettes do not harm the health of users | 428 (69.7) | 186 (30.3) |
| The liquid in e-cigarettes may contain nicotine | 516 (84.0) | 98 (16.0) |
| E-cigarettes do not contain tar | 152 (24.8) | 462 (75.2) |
| The sale of e-cigarettes has been banned throughout Malaysia | 312 (50.8) | 302 (49.2) |
| The use of e-cigarettes is one of a safe way to quit smoking | 388 (63.2) | 226 (36.8) |
| The liquid content in e-cigarettes has been adjusted by the Malaysian Ministry of Health | 223 (36.3) | 391 (63.7) |
| E-cigarette is a new form of addiction in society | 544 (88.6) | 70 (11.4) |
| The use of e-cigarettes has been declared illegal by the National Fatwa Council | 260 (42.3) | 354 (57.7) |

Results of bivariate analyses identified eleven categorical independent variables and ten continuous independent variables with a p-value <0.25. Multiple logistic regression analysis using the “Forward LR” method revealed ten statistically significant factors: sex, race, perceiving that e-cigarettes aid in quitting smoking, perceiving that e-cigarettes do not contain toxic chemicals found in conventional cigarettes, e-cigarette and conventional cigarette usage by close friends, exposure to e-cigarette on television, attitude towards e-cigarettes, willingness to use, and intention to use (see Table 4). These determinants were consistent with the Hosmer and Lemeshow goodness-of-fit test (χ²=11.049, df=8, p=0.199). This model correctly classified 88.4% of e-cigarette use cases.

**Table 4:**
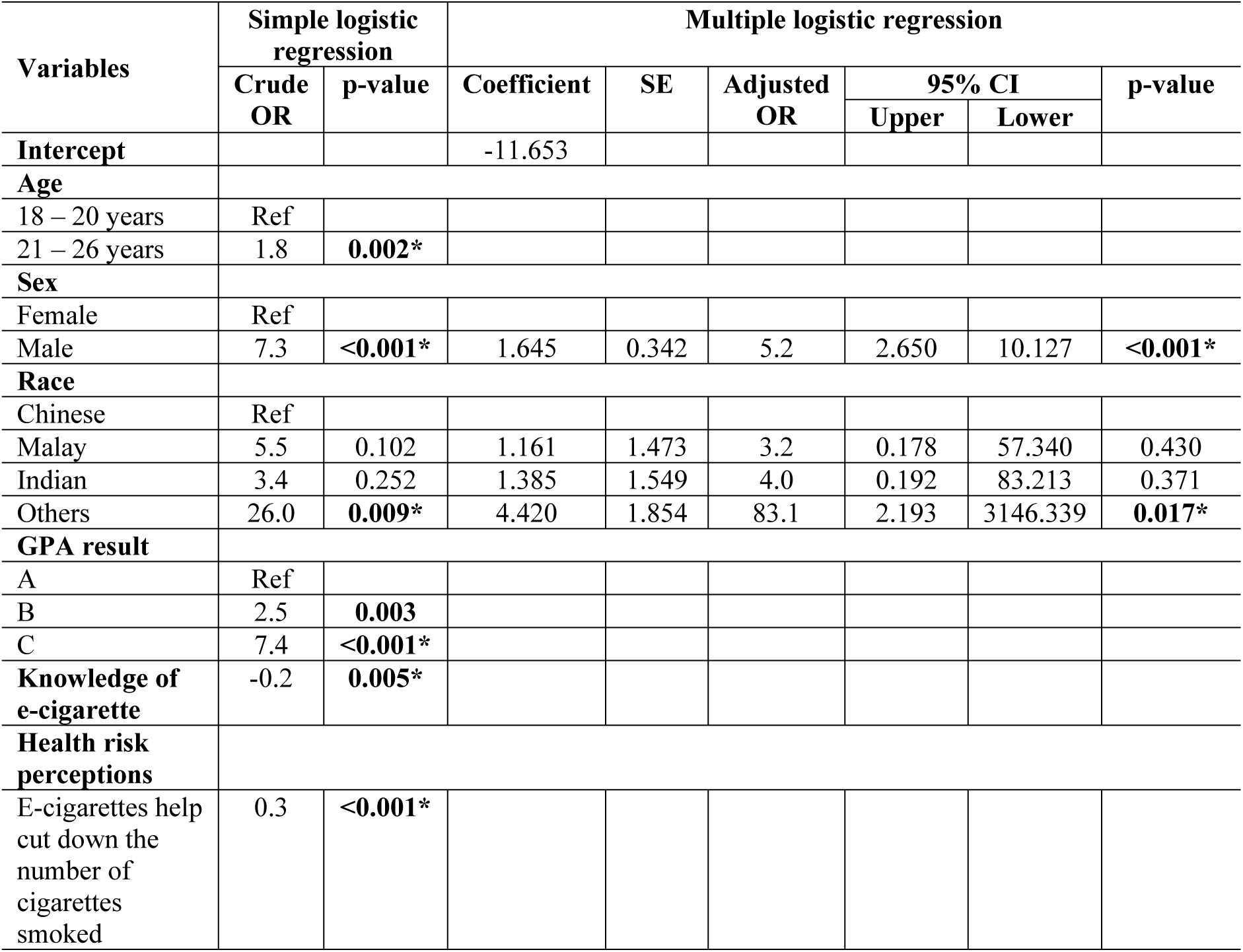
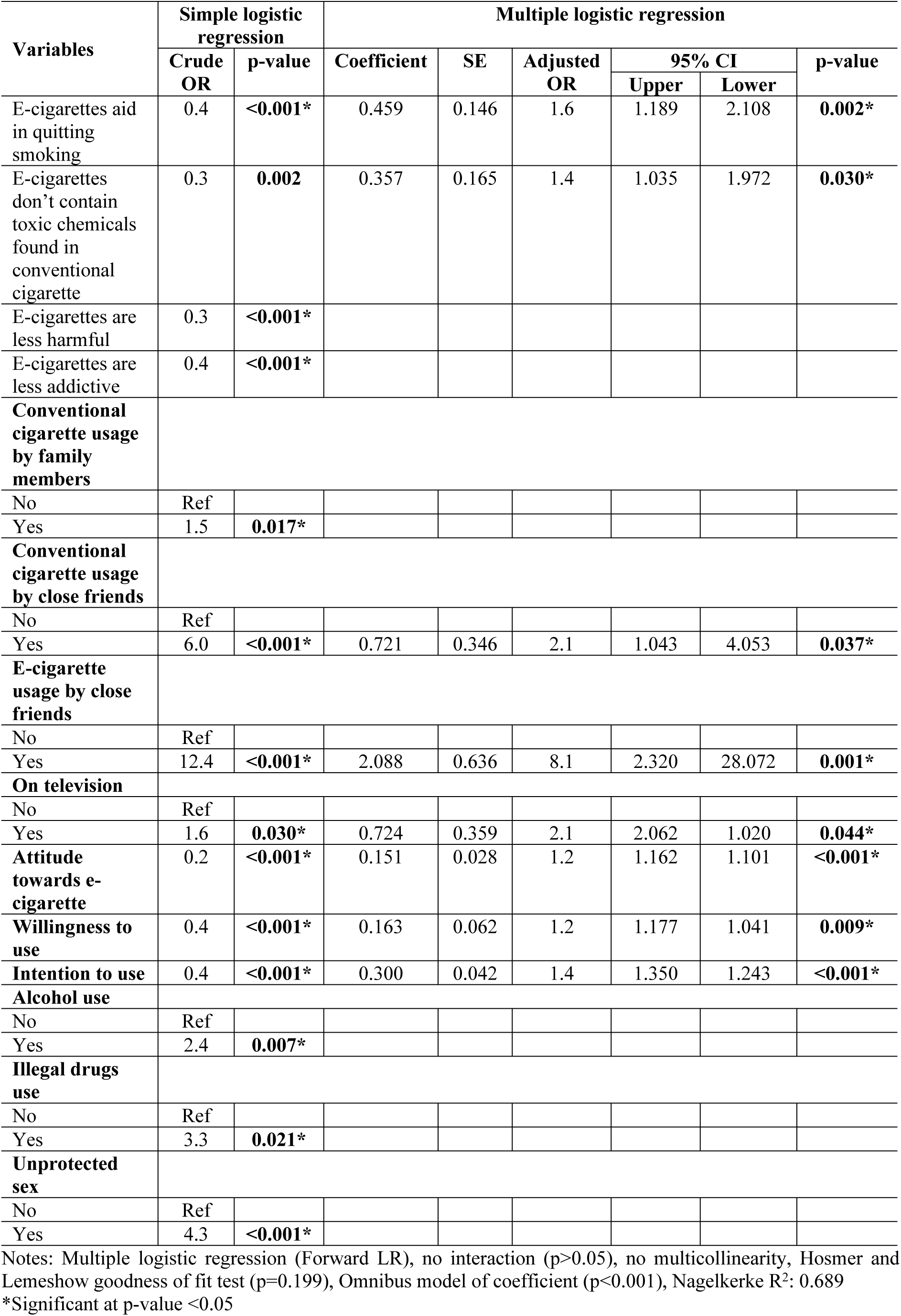
Determinants of e-cigarette use.

| Variables | Simple logistic regression |  | Multiple logistic regression |  |  |  |  |  |
| --- | --- | --- | --- | --- | --- | --- | --- | --- |
|  | Crude OR | p-value | Coefficient | SE | Adjusted OR | 95% CI |  | p-value |
|  |  |  |  |  |  | Upper | Lower |  |
| **Intercept** |  |  | -11.653 |  |  |  |  |  |
| **Age** |  |  |  |  |  |  |  |  |
| 18 – 20 years | Ref |  |  |  |  |  |  |  |
| 21 – 26 years | 1.8 | **0.002\*** |  |  |  |  |  |  |
| **Sex** |  |  |  |  |  |  |  |  |
| Female | Ref |  |  |  |  |  |  |  |
| Male | 7.3 | **<0.001\*** | 1.645 | 0.342 | 5.2 | 2.650 | 10.127 | **<0.001\*** |
| **Race** |  |  |  |  |  |  |  |  |
| Chinese | Ref |  |  |  |  |  |  |  |
| Malay | 5.5 | 0.102 | 1.161 | 1.473 | 3.2 | 0.178 | 57.340 | 0.430 |
| Indian | 3.4 | 0.252 | 1.385 | 1.549 | 4.0 | 0.192 | 83.213 | 0.371 |
| Others | 26.0 | **0.009\*** | 4.420 | 1.854 | 83.1 | 2.193 | 3146.339 | **0.017\*** |
| **GPA result** |  |  |  |  |  |  |  |  |
| A | Ref |  |  |  |  |  |  |  |
| B | 2.5 | **0.003** |  |  |  |  |  |  |
| C | 7.4 | **<0.001\*** |  |  |  |  |  |  |
| **Knowledge of e-cigarette** | -0.2 | **0.005\*** |  |  |  |  |  |  |
| **Health risk perceptions** |  |  |  |  |  |  |  |  |
| E-cigarettes help cut down the number of cigarettes smoked | 0.3 | **<0.001\*** |  |  |  |  |  |  |
|  | Crude OR | p-value | Coefficient | SE | Adjusted OR | 95% CI |  | p-value |
|  |  |  |  |  |  | Upper | Lower |  |
| E-cigarettes aid in quitting smoking | 0.4 | <0.001* | 0.459 | 0.146 | 1.6 | 1.189 | 2.108 | 0.002* |
| E-cigarettes don't contain toxic chemicals found in conventional cigarette | 0.3 | 0.002 | 0.357 | 0.165 | 1.4 | 1.035 | 1.972 | 0.030* |
| E-cigarettes are less harmful | 0.3 | <0.001* |  |  |  |  |  |  |
| E-cigarettes are less addictive | 0.4 | <0.001* |  |  |  |  |  |  |
| **Conventional cigarette usage by family members** |  |  |  |  |  |  |  |  |
| No | Ref |  |  |  |  |  |  |  |
| Yes | 1.5 | 0.017* |  |  |  |  |  |  |
| **Conventional cigarette usage by close friends** |  |  |  |  |  |  |  |  |
| No | Ref |  |  |  |  |  |  |  |
| Yes | 6.0 | <0.001* | 0.721 | 0.346 | 2.1 | 1.043 | 4.053 | 0.037* |
| **E-cigarette usage by close friends** |  |  |  |  |  |  |  |  |
| No | Ref |  |  |  |  |  |  |  |
| Yes | 12.4 | <0.001* | 2.088 | 0.636 | 8.1 | 2.320 | 28.072 | 0.001* |
| **On television** |  |  |  |  |  |  |  |  |
| No | Ref |  |  |  |  |  |  |  |
| Yes | 1.6 | 0.030* | 0.724 | 0.359 | 2.1 | 2.062 | 1.020 | 0.044* |
| **Attitude towards e-cigarette** | 0.2 | <0.001* | 0.151 | 0.028 | 1.2 | 1.162 | 1.101 | <0.001* |
| **Willingness to use** | 0.4 | <0.001* | 0.163 | 0.062 | 1.2 | 1.177 | 1.041 | 0.009* |
| **Intention to use** | 0.4 | <0.001* | 0.300 | 0.042 | 1.4 | 1.350 | 1.243 | <0.001* |
| **Alcohol use** |  |  |  |  |  |  |  |  |
| No | Ref |  |  |  |  |  |  |  |
| Yes | 2.4 | 0.007* |  |  |  |  |  |  |
| **Illegal drugs use** |  |  |  |  |  |  |  |  |
| No | Ref |  |  |  |  |  |  |  |
| Yes | 3.3 | 0.021* |  |  |  |  |  |  |
| **Unprotected sex** |  |  |  |  |  |  |  |  |
| No | Ref |  |  |  |  |  |  |  |
| Yes | 4.3 | <0.001* |  |  |  |  |  |  |

Notes: Multiple logistic regression (Forward LR), no interaction ( $p>0.05$ ), no multicollinearity, Hosmer and Lemeshow goodness of fit test ( $p=0.199$ ), Omnibus model of coefficient ( $p<0.001$ ), Nagelkerke $R^2$ : 0.689
\*Significant at $p$ -value $<0.05$

The model indicates that male respondents were five times more likely to use e-cigarettes compared to females (aOR = 5.2, 95% CI: 2.7-10.1), while respondents from other races (Bumiputera Sabah and Sarawak) were 83 times more likely to use e-cigarettes compared to Chinese (aOR = 83.1, 95% CI: 2.2-3146.3). A one-unit increase in the perception that e-cigarettes aid in quitting smoking was associated with a 1.6-fold increase in the odds of e-cigarette use (aOR = 1.6, 95% CI: 1.2-2.1). Similarly, for each one-unit increase in the perception that e-cigarettes do not contain the toxic chemicals found in conventional cigarettes, the odds of e-cigarette use increased by 1.4 times (aOR = 1.4, 95% CI: 1.0-2.0).

Additionally, having close friends who smoke conventional cigarettes doubled the likelihood of e-cigarette use (aOR = 2.1, 95% CI: 1.0-4.1), while having close friends who use e-cigarettes increased the likelihood by eight times (aOR = 8.0, 95% CI: 2.3-28.1). Exposure to e-cigarette use on television was associated with a twofold increase in e-cigarette use (aOR = 2.1, 95% CI: 1.0-4.2). Moreover, a one-unit increase in positive attitudes towards e-cigarettes was associated with a 1.2-fold increase in the odds of e-cigarette use (aOR = 1.2, 95% CI: 1.1-1.2). Similarly, each one-unit increase in willingness to use and intention to use was linked to a 1.2- fold (aOR = 1.2, 95% CI: 1.0-1.3) and 1.4-fold (aOR = 1.4, 95% CI: 1.2-1.5) increase in the odds of e-cigarette use, respectively. Collectively, these variables accounted for 68.9% of the variance in e-cigarette use among diploma students. However, 31.1% of the variance remains unexplained, indicating the need for further investigation into additional influencing factors.

## Discussion

Our study analyzed e-cigarette use among diploma students at a vocational college in a state in Malaysia. The findings revealed that nearly one in three respondents were e-cigarette users, a prevalence notably higher than that reported in previous research among students in tertiary institutions. For instance, a study among university students in the U.S. aged 18 to 25 years reported an e-cigarette use prevalence of 24.8%, while local studies among university students in Malaysia reported prevalences of 12.4% and 20.4% (2–4). In contrast, a study among students from 12 vocational institutions in Thailand reported a prevalence similar to our current study (28.7%) (6). Overall, the high prevalence of e-cigarette use observed in this study raises significant concerns, particularly among vocational students.

Most respondents were generally aware that e-cigarettes are harmful and represent a new form of addiction in society. However, more than half of the respondents were either unaware of or unsure about the ingredients in e-cigarette liquids, which is consistent with previous study (26). This highlights the critical need for enforcing greater transparency from manufacturers regarding product contents. Some e-cigarette liquids have been found to contain nicotine levels different from those indicated on the label (35). Overall, respondents’ knowledge was rated as moderate, which is lower than the mean score of 5.80±2.04 reported in a previous study (26). Nevertheless, more than half of the respondents were aware of the potential health risks associated with e-cigarette use.

Regarding sex, previous studies have similarly reported a higher likelihood of e-cigarette use among males (5,28,36). In many countries and cultures, including Malaysia, the use of e-cigarettes by males is more socially accepted, whereas females who use e-cigarettes often face cultural disapproval. Female users are frequently perceived as having low moral values and are subject to social ridicule (37). Additionally, males were found to have lower health literacy, seek health information less frequently than females, and are more likely to perceive e-cigarettes as less harmful, making them more prone to use e-cigarettes and engage in other high-risk behaviors (13,38–40).

Our findings showed that individuals from other races were over 80 times more likely to use e-cigarettes compared to the Chinese, while no significant association was observed for the Malay and Indian populations. This disparity may be attributed to specific cultural factors that predispose these groups to higher e-cigarette use, possibly reflecting greater acceptance of the behavior within their social norms. The National Health Morbidity Survey 2019 reported that the prevalence of e-cigarette use was higher among Bumiputera Sabah (5.1%) and Bumiputera Sarawak (5.0%) compared to Chinese (3.5%) (29). A similar finding was observed in a study where white individuals had a higher proportion of e-cigarette use compared to non-whites (χ^2^=8.287, df=2, p=0.016) (31).

Regarding health risk perceptions, our findings are consistent with previous research, which reported a similar association (aOR = 5.197, 95% CI: 2.099-13.106; p<0.001) (25). This perception may arise from the early introduction of e-cigarettes as a supposedly safer alternative to smoking, along with aggressive marketing, claiming that e-cigarettes helped with smoking cessation (13,41). However, e-cigarettes are not a safe alternative, as their use has been linked to significant harm, and some users have become dual users of both e-cigarettes and conventional cigarettes (7,42).

Our findings on the belief that e-cigarettes do not contain the toxic chemicals found in conventional cigarettes are consistent with a previous study, which showed that a significantly higher percentage of e-cigarette users (15.5%) held this perception compared to non-users (7.3%) (p<0.001) (25). The e-cigarette market often promoted e-cigarettes as a healthier alternative to conventional cigarettes. However, recent findings indicate that the chemicals in e-cigarette liquids may cause more harm to users and those around them (15). Additionally, there is a lack of transparency regarding the ingredients and chemicals present in e-cigarette liquids (41,43).

Peer influence was found to be strongly associated with e-cigarette usage in this study. Specifically, the use of conventional cigarettes and e-cigarettes by close friends was found to be one of the factors in this study. Our findings align with previous research, which reported a significant association between having friends using conventional cigarettes and e-cigarette use (OR = 2.72, 95% CI: 1.87-3.95; p<0.001) (21). Similarly, a study in Thailand found a strong association between having friends who use e-cigarettes and e-cigarette usage (aOR = 10.48, 95% CI: 5.96-18.41) (5). This high level of association suggests that e-cigarettes might be increasingly viewed as a socially acceptable or preferred alternative to conventional cigarettes within peer groups. Peer influence significantly impacts individual behavior, especially among young adults, who are more likely to imitate the behaviors of those around them and affect each other’s behavior (44,45). An individual who is offered an e-cigarette by their friends is most likely to accept the offer, which then will lead them to continue using it. They also felt more comfortable within the e-cigarette community (46).

Regarding exposure to e-cigarette use in mass media, such as television, our findings align with a previous U.S. study, which reported that young adults aged 18–24 who were exposed to e-cigarette advertisements on television had a significantly higher likelihood of e-cigarette use compared to other age groups (p<0.05) (47). Television, along with other print media, has been shown to receive the largest share of e-cigarette advertising (48). However, these findings may vary by country, depending on the regulations governing e-cigarette advertising. In Malaysia, the e-cigarette is advertised via print media and social media platforms (41,43). The current trends in social media marketing and advertising should be carefully considered, especially since younger populations are among the highest users of these platforms. Exposure to e-cigarette marketing might have caused individuals to have perception that e-cigarettes are not harmful and do not cause addiction (48).

Another factor identified in this study was attitude. Our finding is consistent with a study conducted in Thailand, which demonstrated that a positive attitude towards e-cigarettes was associated with e-cigarette usage (aOR = 4.171, 95% CI: 2.25-7.734) (6). Additionally, non-smokers were nearly three times more likely to have a negative attitude towards e-cigarettes compared to smokers (aOR = 2.632, 95% CI: 1.725-3.589) (26). An individual’s attitude towards e-cigarettes may be influenced by their knowledge, awareness, and health concerns. Attitude is also shaped by learning from others or through personal experience (49). The significant findings observed in this study may be related to the moderate level of knowledge about e-cigarettes among respondents, which could influence their likelihood of e-cigarette use. This suggests the importance of addressing misconceptions about e-cigarettes and providing balanced information on e-cigarettes as attitudes can be influenced by presenting new and accurate information.

Our finding on the willingness aligns with previous research conducted among male high school students in Iran, which reported that higher willingness to smoke cigarettes was associated with cigarette smoking (OR 1.40, 95% CI: 1.21-1.50, p<0.001) (50). Higher willingness to use e-cigarettes may be influenced by various factors, such as the appeal of flavors, particularly fruit flavors (51). Other than that, decision-making processes in young adults are often more impulsive and less governed by long-term considerations, as they often lack experience compared to older adults, who generally have more experience and better-developed judgment and foresight (52).

Intention is one of the key components studied in the reasoned action pathway of behavioral models, where it has been consistently reported that higher intention is associated with a greater likelihood of performing the behavior. In line with this, this study found a significant association between intention to use and e-cigarette use. Our finding is consistent with a previous study conducted among university students in Saudi Arabia, which found that higher intention was significantly associated with e-cigarette use (AOR 3.5, 95% CI: 2.3-5.3, p<0.001) (53). These findings highlighted the importance of understanding and addressing behavioral intentions in interventions targeting high-risk behaviors. By identifying the factors that influence individuals’ intentions to engage in such behaviors, interventions can be more effectively tailored to promote healthier choices and discourage behaviors associated with adverse health outcomes.

The strengths of this study include a satisfactory response rate of 87.7%, indicating a high level of participant engagement. This study is also the first to examine factors associated with e-cigarette use among students in a vocational college in Malaysia, addressing a significant gap in the literature. Another strength of this study is the sample’s demographic diversity, characterized by an almost equal gender distribution and involvement of students from all diploma courses. This diversity ensures that the findings are representative and generalizable to the broader population of students in tertiary vocational institutions. Additionally, this study provides valuable insights into e-cigarette use within this specific demographic, contributing to the limited research available on this topic in Malaysia. One limitation of the study is that the use of a self-administered questionnaire may have introduced inconsistencies due to social desirability bias or discrepancies between reported and actual experiences. Additionally, there is also limited generalizability of the findings as this study only focused on diploma students from one vocational college and covered a specific time frame.

## Conclusion

In conclusion, this study revealed a notably high prevalence of e-cigarette use among diploma students in a tertiary vocational institution (29.0%), surpassing the rates reported in previous studies. The key determinants for e-cigarette use identified include male gender, Bumiputera Sabah and Sarawak ethnicity, health risk perception (such as beliefs that e-cigarettes aid in quitting smoking and do not contain toxic chemicals found in conventional cigarettes), social influences (such as friends’ use of conventional cigarettes and e-cigarettes), exposure to e-cigarette advertising in media (particularly television), positive attitude towards e-cigarettes, and higher levels of willingness and intention to use e-cigarettes.

It is recommended that further studies be conducted across a more diverse range of vocational institutions in different regions of Malaysia to enhance the generalizability of the findings and to explore potential geographical differences in e-cigarette use among students. Additionally, qualitative research should be conducted, particularly focusing on other ethnic groups (Bumiputera Sabah and Sarawak), to gain a deeper understanding of the factors influencing e-cigarette use in these populations. This approach could refine strategies for preventing e-cigarette use among students. Future studies should also consider incorporating behavioral theory models to provide insights into the underlying motivations and decision-making processes related to e-cigarette use. These findings could significantly contribute to the development of more effective behavioral change interventions targeted at the young adult population.

Health risks associated with e-cigarette use should be included as part of awareness and education programs, given the strong association between health risk perception and e-cigarette usage found in this study. Additionally, information on safe smoking cessation techniques, as well as knowledge about the ingredients and chemicals contained in e-cigarette liquids, should be included in these programs. The current Quit Smoking Program available at registered health clinics, hospitals, and pharmacies should be further promoted and expanded. The current misconception regarding e-cigarette ingredients might have made the public perceive that it is safer than conventional cigarettes.

A ’Quit Vaping’ intervention program may be introduced in institutions for students who use e- cigarettes, potentially including peer support groups. This intervention program also may include promoting healthy behaviors and reducing the normalization of e-cigarette use within social circles, which may benefit the students. As television was identified as one of the key determinants in this study, using this medium to convey e-cigarette awareness and health education could be highly effective. To maximize the impact, collaboration with multiple agencies and stakeholders is essential to ensure comprehensive and accurate content that targets the young adult population. Besides being informative, the content should be creative and interactive, as to attract a younger population. Featuring young adults as the face of the campaign or even as hosts may also enhance its effectiveness

Other than that, enforcing transparency regulations on e-cigarette manufacturers regarding the disclosure of ingredients, particularly nicotine concentrations in e-cigarette liquids, could be a strategic measure to ensure compliance with existing regulations. Strengthening restrictions on e-cigarette advertisements is also crucial. This should include a ban on both implicit and explicit advertisements on television and other platforms popular among young adults. This may help to limit the exposure of e-cigarette promotions to young adults.

## Data Availability

All relevant data are within the manuscript and its Supporting Information files.

## Acknowledgement

The authors would like to express their gratitude to the Ministry of Higher Education for the cooperation and commitment given by the institution and diploma students involved in this study. The authors also would like to thank the Ministry of Higher Education for the permission to publish this article.

## Author Contributions

**Conceptualization**: Siti Munisah Mohd Shoaib, Norliza Ahmad, Aidalina Mahmud

**Performed data collection**: Siti Munisah Mohd Shoaib **Formal analysis**: Siti Munisah Mohd Shoaib, Norliza Ahmad **Supervision**: Norliza Ahmad, Aidalina Mahmud

**Writing – original draft**: Siti Munisah Mohd Shoaib

**Writing – review & editing**: Norliza Ahmad, Aidalina Mahmud

## References

1. Centers for Disease Control & Prevention. About E-Cigarettes (Vapes) [Internet]. 2024 [cited 2024 Aug 30]. Available from: https://www.cdc.gov/tobacco/e-cigarettes/about.html?CDC_AAref_Val=https://www.cdc.gov/tobacco/basic_information/e-cigarettes/about-e-cigarettes.html

2. Wan Puteh SE, Abdul Manap R, Hassan TM, Ahmad IS, Idris IB, Md Sham F, et al. The use of e-cigarettes among university students in Malaysia. Tob Induc Dis [Internet]. 2018 Dec 10 [cited 2024 Aug 30];16(57). Available from: https://www.ncbi.nlm.nih.gov/pmc/articles/PMC6659562/

3. Helmi Ali A, Mohamad Azam IM, Yusof NN, Hussin S. Cross Sectional Study: Cigarette Smoking and the Usage of E-cigarettes Among the Students of University of Cyberjaya. Asian Journal of Medicine and Health Sciences. 2022 Jun;5(1).

4. Jones RD, Asare M, Lanning B. A Retrospective Cross-Sectional Study on the Prevalence of E-cigarette Use Among College Students. J Community Health [Internet]. 2021 Feb 1 [cited 2024 Aug 30];46(1):195–202. Available from: https://www.ncbi.nlm.nih.gov/pmc/articles/PMC7317082/

5. Phetphum C, Prajongjeep A, Thawatchaijareonying K, Wongwuttiyan T, Wongjamnong M, Yossuwan S, et al. Personal and perceptual factors associated with the use of electronic cigarettes among university students in northern Thailand. Tob Induc Dis [Internet]. 2021 Apr 22 [cited 2024 Aug 30];19(31). Available from: https://www.ncbi.nlm.nih.gov/pmc/articles/PMC8059432/

6. Benjakul S, Nakju S, Termsirikulchai L. Factors associated with e-cigarette use among vocational students: A cross-sectional multistage cluster survey, Thailand. Tob Induc Dis [Internet]. 2023 Sep [cited 2024 Aug 30];21(120). Available from: https://www.tobaccoinduceddiseases.org/Factors-associated-with-e-cigarette-use-among-vocational-students-A-cross-sectional,170421,0,2.html

7. Lyzwinski LN, Naslund JA, Miller CJ, Eisenberg MJ. Global youth vaping and respiratory health: epidemiology, interventions, and policies. NPJ Prim Care Respir Med [Internet]. 2022 Apr 11 [cited 2024 Aug 30];32(14). Available from: https://www.ncbi.nlm.nih.gov/pmc/articles/PMC9001701/

8. Centers for Disease Control & Prevention. E-Cigarette, or Vaping, Products Visual Dictionary [Internet]. 2019 [cited 2024 Aug 30]. Available from: https://www.cdc.gov/tobacco/basic_information/e-cigarettes/pdfs/ecigarette-or-vaping-products-visual-dictionary-508.pdf

9. Centers for Disease Control & Prevention. More than 2.5 Million Youth Reported E-Cigarette Use in 2022 [Internet]. 2022 [cited 2024 Aug 30]. Available from: https://www.cdc.gov/media/releases/2022/p1007-e-cigarette-use.html

10. Ministry of Health Malaysia. National Strategic Plan for The Control of Tobacco & Smoking Products 2021-2030 [Internet]. 2021 [cited 2024 Aug 30]. Available from: https://www.moh.gov.my/moh/resources/Penerbitan/Rujukan/NCD/National%20Strategic%20Plan/NCDTembakau20212030.pdf

11. Centers for Disease Control & Prevention. Outbreak of Lung Injury Associated with the Use of E-Cigarette, or Vaping, Product [Internet]. 2020 [cited 2024 Aug 30]. Available from: https://archive.cdc.gov/www_cdc_gov/tobacco/basic_information/e-cigarettes/severe-lung-disease.html

12. Blount BC, Karwowski MP, Shields PG, Morel-Espinosa M, Valentin-Blasini L, Gardner M, et al. Vitamin E Acetate in Bronchoalveolar-Lavage Fluid Associated with EVALI. New England Journal of Medicine [Internet]. 2020 Feb 20 [cited 2024 Aug 30];382(8):697–705. Available from: https://www.ncbi.nlm.nih.gov/pmc/articles/PMC7032996/

13. U.S. Department of Health and Human Services. E-Cigarette Use Among Youth and Young Adults: A Report of the Surgeon General [Internet]. Atlanta, GA: U.S. Department of Health and Human Services, Centers for Disease Control and Prevention, National Center for Chronic Disease Prevention and Health Promotion, Office on Smoking and Health; 2016 [cited 2024 Aug 30]. Available from: https://www.cdc.gov/tobacco/sgr/e-cigarettes/pdfs/2016_sgr_entire_report_508.pdf

14. Almeida-da-Silva CLC, Matshik Dakafay H, O’Brien K, Montierth D, Xiao N, Ojcius DM. Effects of electronic cigarette aerosol exposure on oral and systemic health. Biomed J [Internet]. 2021 Jun [cited 2024 Aug 30];44(3):252–9. Available from: https://www.ncbi.nlm.nih.gov/pmc/articles/PMC8358192/

15. Walley SC, Wilson KM, Winickoff JP, Groner J. A public health crisis: Electronic cigarettes, vape, and JUUL. Pediatrics. 2019 Jun;143(6).

16. Umphres SS, Alarabi AB, Ali HEA, Khasawneh FT, Alshbool FZ. Investigation of the impact of thirdhand e-cigarette exposure on platelet function: A pre-clinical study. Tob Induc Dis [Internet]. 2024 Mar [cited 2024 Aug 30];22(56):1–10. Available from: https://www.ncbi.nlm.nih.gov/pmc/articles/PMC10980912/

17. Tzortzi A, Teloniatis S, Matiampa G, Bakelas G, Tzavara C, Vyzikidou VK, et al. Passive exposure of non-smokers to E-Cigarette aerosols: Sensory irritation, timing and association with volatile organic compounds. Environ Res [Internet]. 2020 Mar [cited 2024 Aug 30];182. Available from: https://www.sciencedirect.com/science/article/pii/S0013935119307601?via%3Dihub

18. Bayly JE, Bernat D, Porter L, Choi K. Secondhand Exposure to Aerosols From Electronic Nicotine Delivery Systems and Asthma Exacerbations Among Youth With Asthma. Chest [Internet]. 2019 Jan [cited 2024 Aug 30];155(1):88–93. Available from: https://www.ncbi.nlm.nih.gov/pmc/articles/PMC6688978/

19. Wipfli H, Bhuiyan MR, Qin X, Gainullina Y, Palaganas E, Jimba M, et al. Tobacco use and E-cigarette regulation: Perspectives of University Students in the Asia-Pacific. Addictive Behaviors [Internet]. 2020 Aug [cited 2024 Aug 30];107. Available from: https://www.sciencedirect.com/science/article/pii/S0306460319310998

20. Cabral P. E-cigarette use and intentions related to psychological distress among cigarette, e-cigarette, and cannabis vape users during the start of the COVID-19 pandemic. BMC Psychol [Internet]. 2022 Dec 1 [cited 2024 Aug 30];10(201). Available from: https://bmcpsychology.biomedcentral.com/articles/10.1186/s40359-022-00910-9#citeas

21. Wang JW, Cao SS, Hu RY. Smoking by family members and friends and electronic-cigarette use in adolescence: A systematic review and meta-analysis [Internet]. Vol. 16, Tobacco Induced Diseases. International Society for the Prevention of Tobacco Induced Diseases; 2018 [cited 2024 Aug 30]. Available from: https://www.ncbi.nlm.nih.gov/pmc/articles/PMC6659504/

22. Coleman M, Donaldson CD, Crano WD, Pike JR, Stacy AW. Associations Between Family and Peer E-Cigarette Use with Adolescent Tobacco and Marijuana Usage: A Longitudinal Path Analytic Approach. Nicotine & Tobacco Research [Internet]. 2021 May [cited 2024 Aug 30];23(5):849–55. Available from: https://www.ncbi.nlm.nih.gov/pmc/articles/PMC8628870/

23. Ali FRM, Dave DM, Colman GJ, Wang X, Saffer H, Marynak KL, et al. Association of e-cigarette advertising with e-cigarette and cigarette use among US adults. Addiction [Internet]. 2021 May 1 [cited 2024 Aug 30];116(5):1212–23. Available from: https://www.ncbi.nlm.nih.gov/pmc/articles/PMC8434873/

24. Chen-Sankey JC, Unger JB, Bansal-Travers M, Niederdeppe J, Bernat E, Choi K. E-cigarette marketing exposure and subsequent experimentation among youth and young adults. Pediatrics [Internet]. 2019 Nov [cited 2024 Aug 30];144(5). Available from: https://www.ncbi.nlm.nih.gov/pmc/articles/PMC6836725/

25. Fauzi R, Areesantichai C. Factors associated with electronic cigarettes use among adolescents in Jakarta, Indonesia. J Health Res [Internet]. 2022 [cited 2024 Aug 30];36(1):2–11. Available from: https://www.emerald.com/insight/content/doi/10.1108/JHR-01-2020-0008/full/pdf?title=factors-associated-with-electronic-cigarettes-use-among-adolescents-in-jakarta-indonesia

26. Jaafar H, Mohd Razi NA, Tengku Mohd TAM, Mohd Noor N ‘Ayn, Mohd Rani MD, Abu Baharin MF, et al. Knowledge, Attitude and Practice on Electronic Cigarette and their Associated Factors among Undergraduate Students in a Public University. IIUM Medical Journal Malaysia [Internet]. 2021 Apr [cited 2024 Aug 30];20(2):43–51. Available from: https://journals.iium.edu.my/kom/index.php/imjm/article/view/506/1178

27. Grant JE, Lust K, Fridberg DJ, King AC, Chamberlain SR. E-Cigarette Use (Vaping) is Associated with Illicit Drug Use, Mental Health Problems, and Impulsivity in University Students. Annals of Clinical Psychiatry [Internet]. 2019 Feb [cited 2024 Aug 30];31(1):27–35. Available from: https://www.ncbi.nlm.nih.gov/pmc/articles/PMC6420081/

28. Seabrook JA, Twynstra J, Gilliland JA. Correlates of Lifetime and Past Month Vape Use in a Sample of Canadian University Students. Subst Abuse [Internet]. 2021 [cited 2024 Aug 30];15. Available from: https://www.ncbi.nlm.nih.gov/pmc/articles/PMC8549468/

29. Institute for Public Health (IPH), National Institutes of Health, Ministry of Health Malaysia. National Health and Morbidity Survey (NHMS) 2019: Vol. I: NCDs – Non-Communicable Diseases: Risk Factors and other Health Problems [Internet]. 2020 [cited 2024 Aug 30]. Available from: https://iku.moh.gov.my/images/IKU/Document/REPORT/NHMS2019/Report_NHMS2019-NCD_v2.pdf

30. Ministry of Higher Education. Statistik Pendidikan Tinggi 2022 - Makro Institusi Pendidikan Tinggi [Internet]. 2023 Jul [cited 2024 Aug 30]. Available from: https://www.mohe.gov.my/en/download/statistics/2022-3/1177-statistik-pendidikan-tinggi-2022-bab-1-makro-institusi-pendidikan-tinggi/file

31. Greer AE, Morgan K, Samuolis J, Diaz G, Merighi J, Mahoney P. An examination of electronic nicotine delivery system use among college students using social cognitive theory. Journal of American College Health. 2022;70(6):1839–47.

32. Vogel EA, Ramo DE, Rubinstein ML, Delucchi KL, Darrow SM, Costello C, et al. Effects of social media on adolescents’ willingness and intention to use e-cigarettes: An experimental investigation. Nicotine and Tobacco Research [Internet]. 2021 Apr 1 [cited 2024 Aug 30];23(4):694–701. Available from: https://www.ncbi.nlm.nih.gov/pmc/articles/PMC7976937/

33. Tavakol M, Dennick R. Making sense of Cronbach’s alpha. Int J Med Educ [Internet]. 2011 [cited 2024 Aug 30];2:53–5. Available from: https://www.ncbi.nlm.nih.gov/pmc/articles/PMC4205511/

34. Hosmer DW, Lemeshow S. Applied Logistic Regression. Second. JohnWiley&Sons,Inc; 2000.

35. Bozier J, Chivers EK, Chapman DG, Larcombe AN, Bastian NA, Masso-Silva JA, et al. The Evolving Landscape of e-Cigarettes: A Systematic Review of Recent Evidence. Chest. 2020 May 1;157(5):1362–90.

36. Ahmed LA, Verlinden M, Alobeidli MA, Alahbabi RH, Alkatheeri R, Saddik B, et al. Patterns of tobacco smoking and nicotine vaping among university students in the united arab emirates: A cross-sectional study. Int J Environ Res Public Health. 2021 Jul 19;18(14).

37. Indriani RC, Aulia F, Sianturi PA, Sundari P, Sembiring SUB, Milala YY, et al. The Phenomenon of Vaping in Female Students. Indonesian Journal of Medical Anthropology [Internet]. 2023 Oct 3 [cited 2024 Aug 30];4(2):44–9. Available from: https://www.researchgate.net/publication/374423754_The_Phenomenon_of_Vaping_i n_Female_Students#fullTextFileContent

38. Svendsen MT, Bak CK, Sørensen K, Pelikan J, Riddersholm SJ, Skals RK, et al. Associations of health literacy with socioeconomic position, health risk behavior, and health status: A large national population-based survey among Danish adults. BMC Public Health [Internet]. 2020 Apr 28 [cited 2024 Aug 30];20(1). Available from: https://www.ncbi.nlm.nih.gov/pmc/articles/PMC7187482/

39. Hassan S, Masoud O. Online health information seeking and health literacy among non-medical college students: gender differences. J Public Health (Bangkok). 2021 Dec 1;29(6):1267–73.

40. Martinović I, Un Kim S, Stanarević Katavić S. Study of health information needs among adolescents in Croatia shows distinct gender differences in information seeking behaviour. Health Info Libr J. 2023 Mar 1;40(1):70–91.

41. van der Eijk Y, Tan Ping Ping G, Ong SE, Tan Li Xin G, Li D, Zhang D, et al. E-Cigarette Markets and Policy Responses in Southeast Asia: A Scoping Review. Int J Health Policy Manag [Internet]. 2022 Sep 1 [cited 2024 Aug 30];11(9):1616–24. Available from: https://www.ncbi.nlm.nih.gov/pmc/articles/PMC9808234/

42. Burrowes KS, Fuge C, Murray T, Amos J, Pitama S, Beckert L. An evaluation of a New Zealand “vape to quit smoking” programme. New Zealand Medical Journal [Internet]. 2022 Aug 19 [cited 2024 Aug 30];135(1560):45–55. Available from: https://www.researchgate.net/publication/363231669_An_evaluation_of_a_New_Zeal and_vape_to_quit_smoking_programme

43. Shroff SM, Sreeramareddy CT. Marketing claims, promotional strategies, and product information on Malaysian e-cigarette retailer websites-a content analysis. Subst Abuse Treatment, Prevention, and Policy [Internet]. 2024 [cited 2024 Aug 30];19(1). Available from: https://www.ncbi.nlm.nih.gov/pmc/articles/PMC10809498/

44. Addo IY, Acquah E, Nyarko SH, Dickson KS, Boateng ENK, Ayebeng C. Exposure to pro-tobacco and anti-tobacco media messages and events and smoking behaviour among adolescents in Gambia. BMC Public Health [Internet]. 2024 Apr 15 [cited 2024 Aug 30];24. Available from: https://bmcpublichealth.biomedcentral.com/articles/10.1186/s12889-024-18543-5

45. Arduini-Van Hoose N. Adolescent Psychology [Internet]. 2020 [cited 2024 Feb 13]. Available from: https://adolescentpsychology.pressbooks.sunycreate.cloud/

46. Langley T, Bell-Williams R, Pattinson J, Britton J, Bains M. ‘I felt welcomed in like they’re a little family in there, i felt like i was joining a team or something’: Vape shop customers’ experiences of E-cigarette use, vape shops and the vaping community. Int J Environ Res Public Health [Internet]. 2019 Jul 1 [cited 2024 Aug 30];16(13). Available from: https://www.ncbi.nlm.nih.gov/pmc/articles/PMC6652145/

47. Ali FRM, Dave DM, Colman GJ, Wang X, Saffer H, Marynak KL, et al. Association of e-cigarette advertising with e-cigarette and cigarette use among US adults. Addiction. 2021 May 1;116(5):1212–23.

48. Collins L, Glasser AM, Abudayyeh H, Pearson JL, Villanti AC. E-cigarette marketing and communication: How E-Cigarette Companies Market E-Cigarettes and the Public Engages with E-cigarette Information. Nicotine and Tobacco Research [Internet]. 2019 Jan 1 [cited 2024 Aug 30];21(1):14–24. Available from: https://www.ncbi.nlm.nih.gov/pmc/articles/PMC6610165/

49. Pickens J. Attitudes and Perceptions [Internet]. Borkowski N, editor. Organizational Behavior in Health Care. Sudbury,Massachusetts: Jones and Bartlett Publishers; 2005 [cited 2024 Aug 30]. 43–75 p. Available from: https://www.researchgate.net/publication/267362543_Attitudes_and_Perceptions#fullTextFileContent

50. Farshidi H, Aghamolaei T, Hosseini Z, Nejad AG, Hosseini FA. Cigarette smoking based on prototype willingness model in male high school students. Int J High Risk Behav Addict [Internet]. 2017 Nov 1 [cited 2024 Aug 30];7(1). Available from: https://brieflands.com/articles/ijhrba-63209

51. Abadi MH, Lipperman-Kreda S, Shamblen SR, Thompson K, Grube JW, Leventhal AM, et al. The impact of flavored ENDS use among adolescents on daily use occasions and number of puffs, and next day intentions and willingness to vape. Addictive Behaviors [Internet]. 2021 Mar 1 [cited 2024 Aug 30];114. Available from: https://www.ncbi.nlm.nih.gov/pmc/articles/PMC7785609/

52. Barati M, Allahverdipour H, Hidarnia A, Niknami S. Predicting Tobacco Smoking Among Male Adolescents in Hamadan City, West of Iran in 2014: An Application of the Prototype Willingness Model. J Res Health Sci [Internet]. 2015 [cited 2024 Aug 30];15(2):113–8. Available from: https://www.researchgate.net/publication/280123218_Predicting_Tobacco_Smoking_Among_Male_Adolescents_in_Hamadan_City_West_of_Iran_in_2014_An_Application_of_the_Prototype_Willingness_Model

53. Alduraywish SA, Aldakheel FM, Alsuhaibani OS, Jabaan AD, Alballa RS, Alrashed AW, et al. Knowledge and Attitude toward E-Cigarettes among First Year University Students in Riyadh, Saudi Arabia. Healthcare [Internet]. 2023 Feb 1 [cited 2024 Aug 30];11(4). Available from: https://www.ncbi.nlm.nih.gov/pmc/articles/PMC9957237/

